# Privacy Protection of Sexually Transmitted Infections Information from Chinese Electronic Medical Records

**DOI:** 10.1101/2024.08.13.24311908

**Authors:** Mengchun Gong, Yue Yu, Zihao Ouyang, Wenzhao Shi, Chao Liu, Qilin Wang, Jiale Nan, Endi Cai, Fen Ding, Sheng Nie

**Author notes:** Corresponding Author: Mengchun Gong, Sheng Nie. These authors contributed equally (Mengchun Gong, Yue Yu, Zihao Ouyang).

## Abstract

**Objectives:** To formulate an efficacious approach for safeguarding the privacy information of electronic medical records.

**Design:** Chinese patient electronic medical record text information.

**Setting:** The Chinese Renal Disease Data System database.

**Participants:** 3,233,174 patients between 1 Jan. 2010 and 31 Dec. 2023.

**Main outcome measures:** Annotated patient privacy fields and the effectiveness of privacy protection

**Results:** We have developed an automated tool named EPSTII, designed to protect the privacy of patients’ sexually transmitted infection information within medical records. Through the refinement of keywords and the integration of expert knowledge, EPSTII currently achieves a 100% accuracy and recall rate. Our privacy protection measures have reached a 99.5% success rate, ensuring the utmost protection of STI patients’ privacy. As the first large-scale investigation into privacy leakage and STI identification in Chinese electronic medical records, our research paves the way for the future development of patient privacy protection laws in China and the advancement of more sophisticated tools.

**Conclusions:** The EPSTII method demonstrates a feasible and effective approach to protect privacy in electronic medical records from 19 hospitals, offering comprehensive insights for infectious disease research using Chinese electronic medical records, with protocols tailored for accurate STI data extraction and enhanced protection compared to traditional methods.

## Introduction

The comprehensive adoption of electronic medical records (EMRs) offers both tangible and intangible benefits to hospitals,^1^ including enhanced quality of care, reduced medical errors, decreased costs,^2^ and unrestricted access to patient information across time and space.^3^ However, medical information is generally considered highly sensitive, and any privacy breach can cause direct or indirect harm to patients.^4^ The increasing reliance on EMRs correspondingly raises the potential negative impact of privacy breaches through EMRs, involving prescription records,^5^ diagnostic codes,^6^ genomic data with allele frequency,^7^ and improperly published medical data. These breaches can lead to unintended losses for both hospitals and patients. Such privacy breaches can damage an individual’s social reputation and normal social behavior and can be nearly destructive for patients with sexually transmitted infections (STIs).

Personal information related to STIs is directly tied to personal privacy and personality rights. Protecting this information is essential for safeguarding individual human rights. Studies have shown that if hospitals lack adequate protective and management measures, these privacy concerns can become public knowledge through word of mouth, causing significant psychological harm to patients and deterring them from seeking standardized treatment.^8,9^ Moreover, due to the particular nature of STIs, personal information also involves the legitimate interests of others and public health safety.^10.11^ International experience demonstrates that focusing unilaterally on either aspect can lead to mistakes.^12^ Overemphasizing public health safety and the unilateral and mandatory nature of STI information management may infringe upon the privacy rights of the individual, while overemphasizing the absolute protection of individual privacy rights may harm the legitimate interests of others (such as the health and life rights of their sexual partners) and public health safety (such as accelerating the spread of AIDS). The correct approach is to properly balance the protection and management of personal information related to STIs in all relevant fields, to achieve mutual reinforcement and promotion of health and human rights.^13^

The fear of disclosure and its associated consequences is particularly pronounced in certain cultural contexts. When deciding to access HIV counseling, testing, and treatment services, clients often worry about the potential for their status to be disclosed and the negative consequences or risks that may follow. Lyimo et al pointed out that these risks are associated with societal beliefs and perceptions of the disease,^14^ as HIV/AIDS is viewed as contagious, severe, life-threatening, and potentially caused by norm-violating behaviors, such as prostitution, homosexuality, and promiscuity. In Ghana, these risks are often culturally perceived as shameful (animguaseè), disrespectful (onnibuo), and dishonorable (onnianimuonyam). Consequences include divorce, rejection, ostracism, discrimination, and unemployment.^15,16^ Fear of stigmatization and its consequences negatively impacts potential clients seeking HIV testing, diagnosis, and treatment in sub-Saharan African countries, including Ghana. This situation poses a serious challenge for stakeholders committed to preventing the spread of HIV/AIDS and infections like that.^17^

Currently, due to the lack of robust management policies and technical safeguards for privacy protection in Chinese EMRs, and the absence of a unified industry standard, unauthorized use, leakage, and even illegal selling of medical data and information are becoming rampant.^18^ With the continuous increase in patients’ self-protection awareness, medical disputes arising from inadequate protection of sensitive information have surged, significantly affecting the process of building an effective privacy protection mechanism. Therefore, addressing these issues has become one of the primary challenges in promoting the collection, transmission, and sharing of Chinese EMR data.

According to the “Technical Specification of Hospital Information Platform Based on Electronic Medical Records” released by the National Health Commission of China in 2014, different health institutions nationwide share a similar EMR framework. This framework encompasses sub-datasets including diagnostic information, medical advice, laboratory test results, examination information, and surgical records.^19^ Despite this standardization, issues with the continuity and integrity of EMR documentation present significant challenges for multicenter data integration.^20^ Traditional strategies for information extraction and privacy protection in Real-World Evidence (RWE) research and clinical data transfer typically focus on fixed sub-datasets, such as diagnosis and present illness history, as well as direct data entities, such as confirmed records in patient EMRs. These approaches, known as fixed location identification strategies, result in biases in patient inclusion and inadequacies in data shielding in RWE research.

This study aims to develop a protocol for the automatic and precise extraction of STI information from Chinese EMRs with the highest possible accuracy. This information is vital for patient inclusion and cohort identification in RWE research and for addressing the issue of STI information leakage.^21,22^ Additionally, this study will formulate privacy protection strategies to optimally safeguard STI data and identify privacy leakage risks within different sub-datasets of EMRs. To our knowledge, this research is the first to quantify the frequency of STI privacy information in Chinese EMRs, thereby enabling consideration of associated risks when utilizing patient EMRs for RWE research.

## Method

### Data Source

This retrospective study utilized the Chinese Renal Disease Data System (CRDS) database, a comprehensive national EMR repository. The CRDS comprises data from 19 tertiary referral hospitals across 10 provinces, representing China’s five geographical regions: North, Central, East, South, and Southwest. Complete EMRs from each hospital were transferred to the central database at Nanfang Hospital of Southern Medical University in Guangzhou. This database includes the EMRs of all patients who visited between the beginning of 1995 and the end of 2023, encompassing a total of 11,759,139 patients.

### Structure of Chinese EMRs

According to Chinese national specifications for standard EMR structure, Chinese EMRs consist of similar sub-datasets with minor differences in nomenclature, including diagnosis, test results, examine results, surgical sheets, medication, and medical record texts. The medical record texts are further divided into ten sections: course records, admission records, discharge records, referral records, consultation records, nursing records, death records, surgical notes, informed consent forms, and others. In the CRDS, the admission record texts have been preprocessed using natural language processing (NLP) to include allergic history, chief complaint, disease history, tobacco and alcohol history, family history, marriage history, surgical history, and toxic exposure history. The general structure of Chinese EMRs is illustrated in Figure 1.

**Figure 1.**
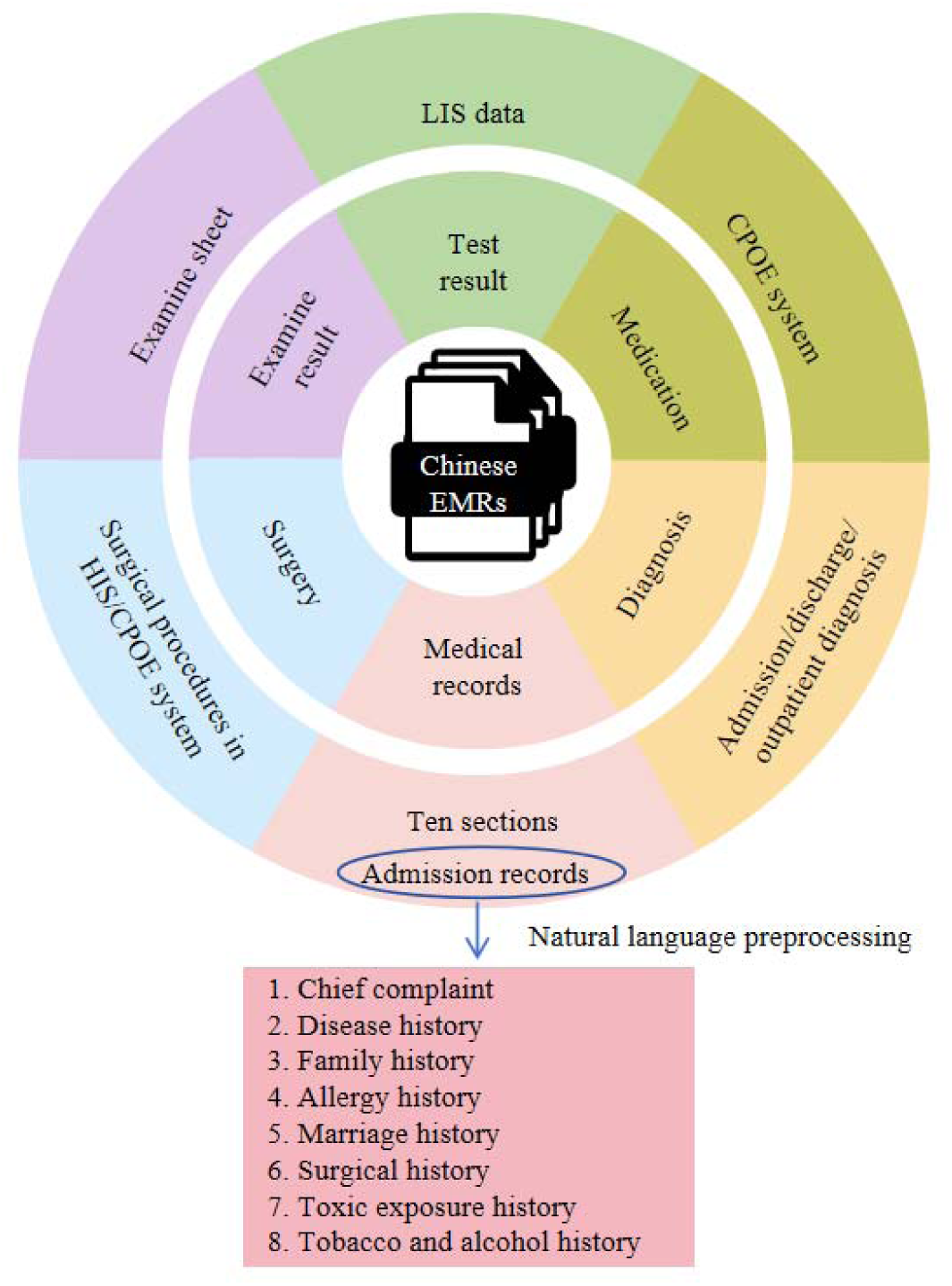
Structure of Chinese EMRs. The EMRs consist of medical records, surgery, exam and test results, medication, and diagnosis. The admission records had been preprocessed by NLP. LIS: laboratory information system. HIS: hospital information system. CPOE: computerized physician order entry.

### Patient Inclusion

This study utilized data from the CRDS database, encompassing patient records from January 1, 2010 to December 31, 2020. The statistical timeline included each patient’s last visit information, along with all previous medical histories. After removing duplicate entries, a random selection of 50% of the patients was made for statistical analysis. The admission record texts were preprocessed using NLP techniques within the CRDS to extract details and improve readability.

### Enhance the Dictionary of STI Utilizing Public Corpora

The key term extraction in this paper utilizes the word2vec method from the field of NLP.^23^ Initially, the names of STIs, along with their synonyms, are used as entry words for retrieval. Searches are conducted on internet diagnostic and treatment platforms, as well as online medical Q&A platforms, to acquire textual information about the diseases, covering dimensions such as diagnosis, related surgical procedures, relevant chief complaint descriptions, history of STIs, and descriptions of STIs.

The textual information for each disease is integrated into a single document, and “cut_all” word segmentation strategy is performed using the Jieba tool to incorporate as many word combinations as possible, while avoiding situations where words nested within a longer word cannot be recognized. For long terms (usually with more than 5 characters) that cannot be accurately segmented, a domain-specific dictionary composed of disease-related terms and vocabulary is formulated to assist Jieba segmentation seeing Figure 2.

**Figure 2.**
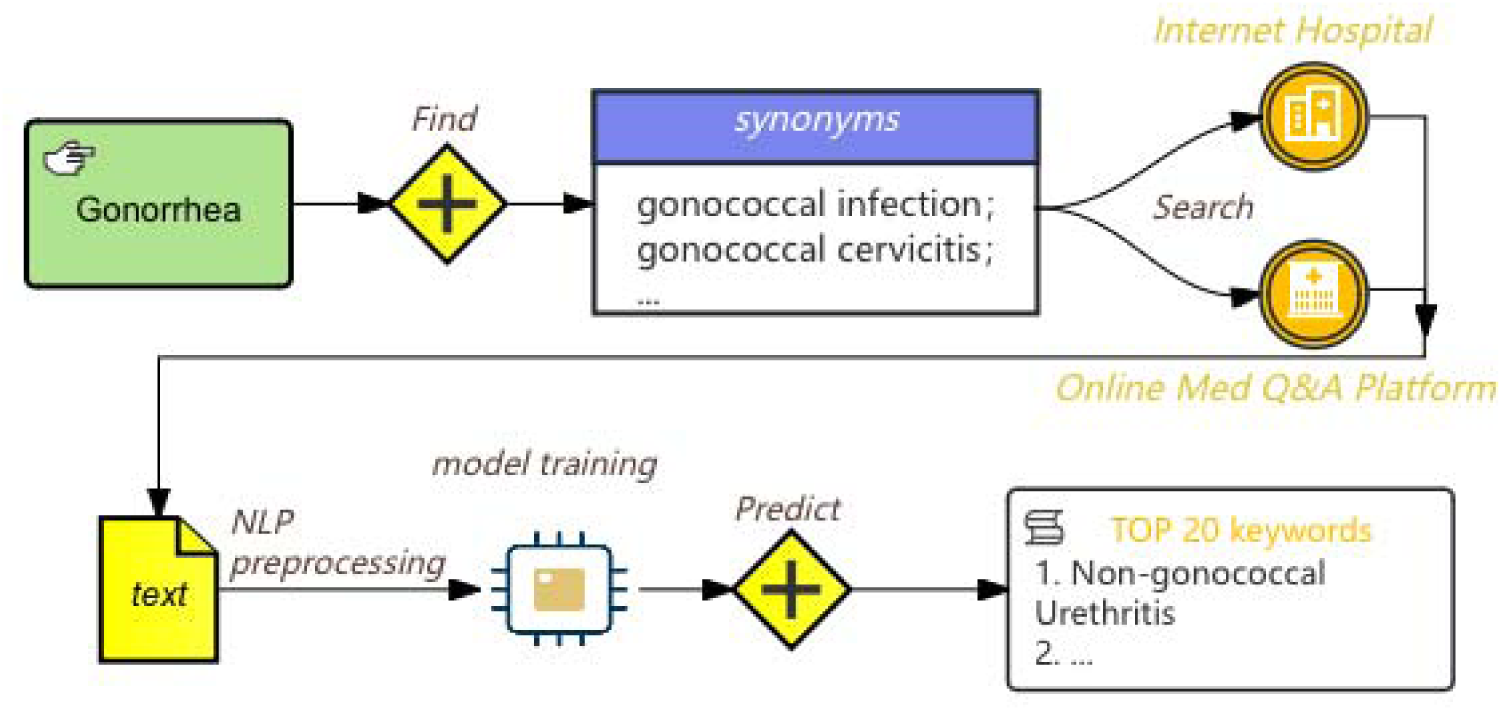
Flowchart of keywords generation by word2vec. Gonorrhea, including its synonyms, were used to search online to collect information about the disease. NLP method was used to analyze the text file and obtain 20 most relevant keywords.

After obtaining the segmentation results, the word2vec method is used to vectorize the text representation. We adopt a grid search strategy in training the word2vec model to obtain the optimal information representation.^24^ Once training is completed, we use the model to predict each disease’s top 20 most relevant keywords to fulfill the dictionary, and the more index value closer to 1, the more relevant they are. The detail hyperparameter settings for the two methods are in supplementary table 1.

### Extract STI Information Using Regular Expressions

The research team conducted preliminary investigations on Chinese EMRs and, based on the aforementioned dictionary, developed the Extraction Protocol of Sexually Transmitted Infections Information (EPSTII) with input from experts, textbooks, guidelines, and relevant literature. These rules were repeatedly refined, adjusted, verified, and continuously improved, taking into account the characteristics of medical record writing in various hospitals. Traditional methods typically extract patient data from the Chinese EMR diagnosis sheet using diagnostic coding. In contrast, we initially established identification rules for detection results.

Given the diversity and complexity of the medical coding system in Chinese EMRs, we employed regular expressions (regex) to search for STI information across the entire EMR, rather than relying solely on diagnostic codes in specific sub-datasets. All regex search patterns were developed through expert meetings and discussions.

### Manual Curation and Verification

Initially, we utilized the EPSTII to extract patients with STI information from the disease history database comprising 1,634,877 patients. Next, we randomly selected 1,000 patients from those with STI information and another 1,000 patients from those without STI information, both drawn from the same disease history database. Under the guidance of two experts, we calculated the precision and recall rates using the formulas provided below.

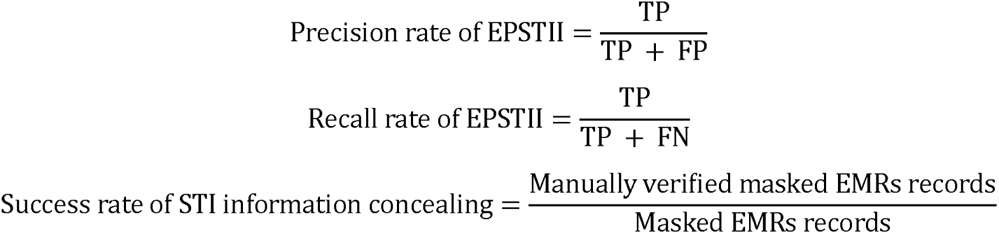

True Positives (TP) were patients correctly identified as having STI information, while False Positives (FP) were patients incorrectly identified as having STI information. True Negatives (TN) were patients correctly identified as not having STI information, and False Negatives (FN) were patients incorrectly identified as not having STI information. Precision, the ratio of TP to the total predicted positives (TP + FP), indicates the accuracy of positive predictions. Recall, the ratio of TP to all actual positives (TP + FN), measures the model’s ability to identify actual positive cases.

After continuously refining the EPSTII rules to achieve higher precision and recall rates, we applied the EPSTII rules to the remaining EMR to extract patients with STI information.

### Privacy Protection Strategies

Based on the analysis of the located information, we have developed privacy protection strategies to prevent the unnecessary and inadvertent exposure of sensitive data during the RWE analysis process. Due to the diverse writing styles found in medical records, inadequate de-identification may leave some sensitive patient information unprotected, while excessive de-identification may obscure other relevant data. Initially, the EPSTII was used to identify keywords related to STIs. With expert guidance, we decided to replace each of these keywords, along with the 10 characters before and after them, with asterisks (*) to de-identify sensitive STI information, thereby safeguarding patient privacy. This approach minimizes the risk of inferring patients’ STI information from EMRs.

Reviewers attempted to identify patients with STIs within the privacy-masked sample to test the success rate of the privacy protection strategies, where the formula is shown above.

When it is unavoidable to use information related to the STI, we systematically estimated the frequency of patient identity and privacy information use to assess the risk of unnecessary privacy exposure.

### Ethical Considerations

This study received approval from the Medical Ethics Committee of Nanfang Hospital, Southern Medical University (approval number: NFEC-2019-213), with a waiver for patient informed consent due to its retrospective design. Additionally, it was approved by the China Office of Human Genetic Resources for Data Preservation Application (approval number: 2021-BC0037).

## Result

### Patient Inclusion

After removing duplicate entries and applying a 50% entry ratio, 3,233,174 patients from various age groups who visited between January 1, 2010 and December 31, 2020 were selected. Notably, removing duplicates reduced the sample size from 10,539,648 to 6,466,357 due to multiple diagnostic records for individual patients. Among the sample, 3,102,028 patients had diagnosis records and 1,735,799 patients had complete medical records. The detail distribution of the EMR sub-datasets and the workflow of our study is shown in Figure 3.

**Figure 3.**
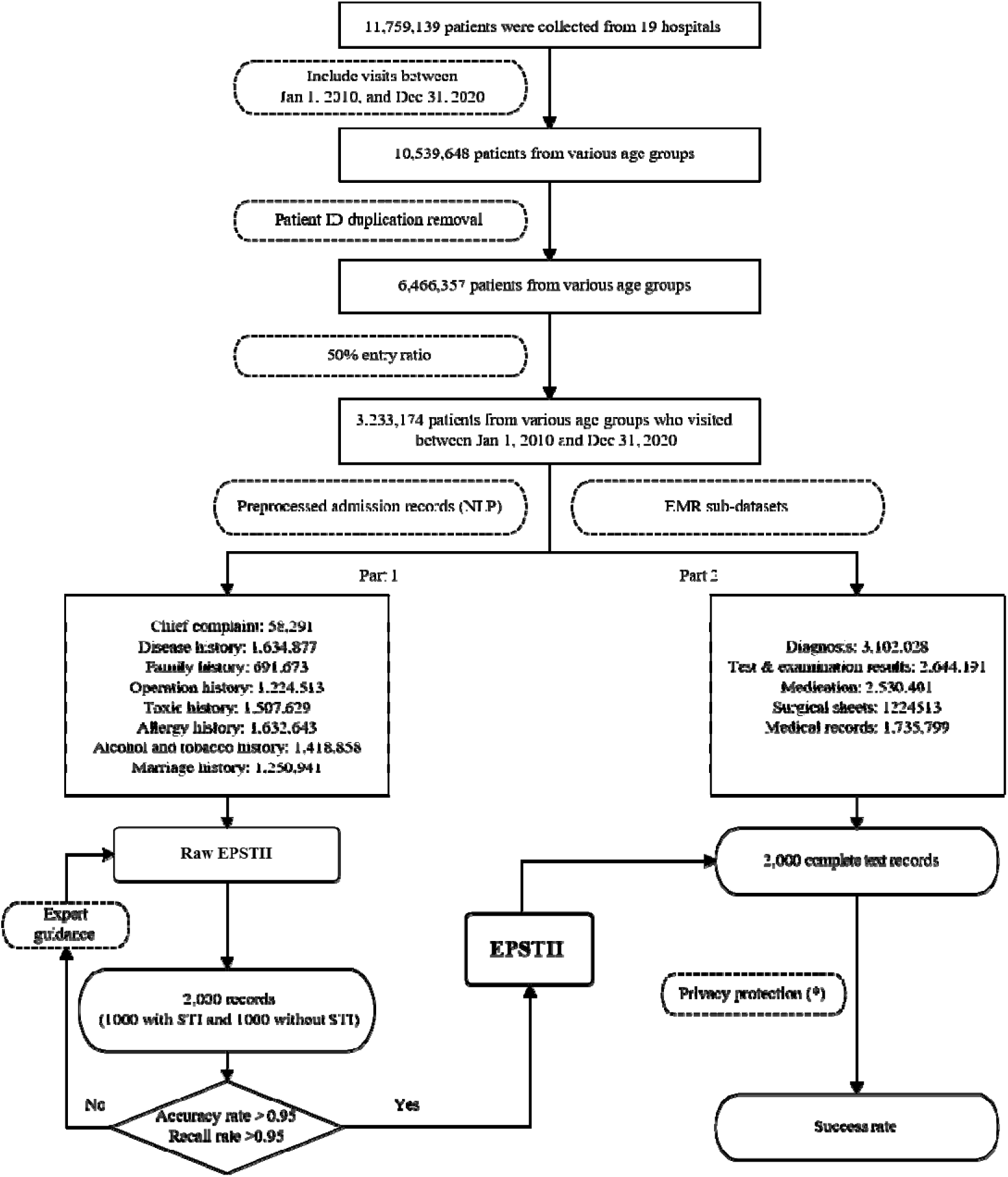
Workflow chart. The initial database had 11,759,139 patients. After selecting time span, removing duplicates, and applying a 50% entry ratio, the final sample consisted of 3,233,174 observations. Admission records were preprocessed by NLP resulting in 8 sub-datasets. EPSTII was used to extract patients with STI information. 1000 patients with STI information and 1000 patients without STI information were randomly selected to calculate precision and recall rate. Expert guidance helped refine the EPSTII rules. Once the protocol performed satisfactorily, EPSTII was applyed to the 2000 randomly selected EMRs to calculate the success rate of privacy protection strategy.

### Regex Search Pattern

After using the grid search method to train the word2vec model, the following keywords are obtained (Table 1. Taking gonorrhea and syphilis as examples, more details can be found in the Supplementary Table 2).

**Table 1.** Top 20 keywords generated by word2vec. The 3 columns are English translation, original Chinese text, and its relativity index.

| Syphilis |  |  |  |  |
| --- | --- | --- | --- | --- |
| Order | Related Keywords | Relativity Index(0~1) | Related Keywords | Relativity Index(0~1) |
| 1 | Non-gonococcal Urethritis | 0.691 | Dark Field | 0.638 |
| 2 | Urethritis | 0.638 | Exclude | 0.636 |
| 3 | Symptom | 0.636 | Spirochete | 0.633 |
| 4 | gonococcus | 0.608 | HSV | 0.607 |
| 5 | General Term | 0.566 | Ulcer | 0.573 |
| 6 | High Magnification | 0.559 | Insidious | 0.571 |
| 7 | Field | 0.550 | Identify | 0.566 |
| 8 | Secretion | 0.536 | Barber | 0.558 |
| 9 | Light(adj.) | 0.531 | Soft Chancre | 0.555 |
| 10 | Quite A Little | 0.528 | Genital Herpes | 0.552 |
| 11 | Oral Sex | 0.497 | (meaningless) | 0.551 |
| 12 | Chlamydia | 0.496 | Inguen | 0.544 |
| 13 | Mucus | 0.496 | (meaningless) | 0.529 |
| 14 | First | 0.496 | Capsule | 0.527 |
| 15 | Urine | 0.496 | Lymph Node | 0.524 |
| 16 | Practicing Safe Sex | 0.495 | Exude | 0.514 |
| 17 | Inflammation | 0.494 | Serology | 0.510 |
| 18 | Fungus | 0.494 | Multiple | 0.510 |
| 19 | Teenager | 0.493 | Chancre | 0.509 |
| 20 | Outflow | 0.493 | Bacillus | 0.504 |

Based on the above dictionary, we investigated Chinese EMRs and, incorporating input from experts, textbooks, guidelines, and relevant literature, developed the EPSTII. Given the unique characteristics of medical record writing in various hospitals, these rules underwent iterative refinement, adjustment, validation, and continuous improvement. All regular expression search patterns were developed through expert meetings and discussions.

To implement regex, we utilized R software (version 4.2.2) to extract STI privacy information from each sub-dataset. In the following example, “disease_history” represents the sub-dataset of preprocessed admission records, and “disease_name” is the field containing the name of the disease. STI_privacy has all the patients’ records that have the disease in the search patterns.

STI_privacy = disease_history[grep(disease_history$disease_name, fixed = FALSE)]

The regular expression rules enable us to determine the proportion of patients with a history of STIs across various sub-datasets of the EMR and to quantify the frequency of STI information.

### Number of STI Information

Following preliminary investigations, we applied the EPSTII to the EMRs of 3,233,174 patients. Table 2 presents the distribution of STI information in diagnosis records from the 19 hospitals.

**Table 2.** Distribution of patients with STI information in 19 hospitals. The 4 columns are city area, total number of patients, total patients with STI information, and their proportions.

| Hospita |  |  | Total patients with | Percentage |
| --- | --- | --- | --- | --- |
| I | City and Area | Total patients | STI | (%) |
| 1 | Guangzhou, Southern | 271715 | 31,676 | 11.66 |
| 2 | Beijing, Northern | 90901 | 2,038 | 2.24 |
| 3 | Jinan, Northern | 269813 | 5,116 | 1.90 |
| 4 | Hangzhou, Eastern | 248390 | 6,928 | 2.79 |
| 5 | Hangzhou, Eastern | 192342 | 7,089 | 3.69 |
| 6 | Guangzhou, Southern | 176901 | 3,780 | 2.14 |
| 7 | Shenzhen, Southern | 100351 | 2,837 | 2.83 |
| 8 | Nanjing, Eastern | 149751 | 520 | 0.35 |
| 9 | Shanghai, Eastern | 96822 | 1,822 | 1.88 |
| 10 | Chengdu,<br>Southwestern | 228432 | 8,444 | 3.70 |
| 11 | Hefei, Eastern | 316766 | 11,015 | 3.48 |
| 12 | Wuhan, Central | 20024 | 1,359 | 6.79 |
| 13 | Maoming, Southern | 204203 | 11,455 | 5.61 |
| 14 | Guangzhou, Southern | 180998 | 9,637 | 5.32 |
| 15 | Huizhou, Southern | 68694 | 6,659 | 9.69 |
| 16 | Guiyang,<br>Southwestern | 45654 | 449 | 0.98 |
| 17 | Foshan, Southern | 267341 | 22,741 | 8.51 |
| 18 | Guangzhou, Southern | 79833 | 9,623 | 12.05 |
| 19 | Guangzhou, Southern | 93097 | 5,668 | 6.09 |
| SUM |  | 3102028 | 148,856 |  |

We also plot the distribution and proportion of patients with STI in each year, as shown in Figure 4 and Figure 5.

**Figure 4.**
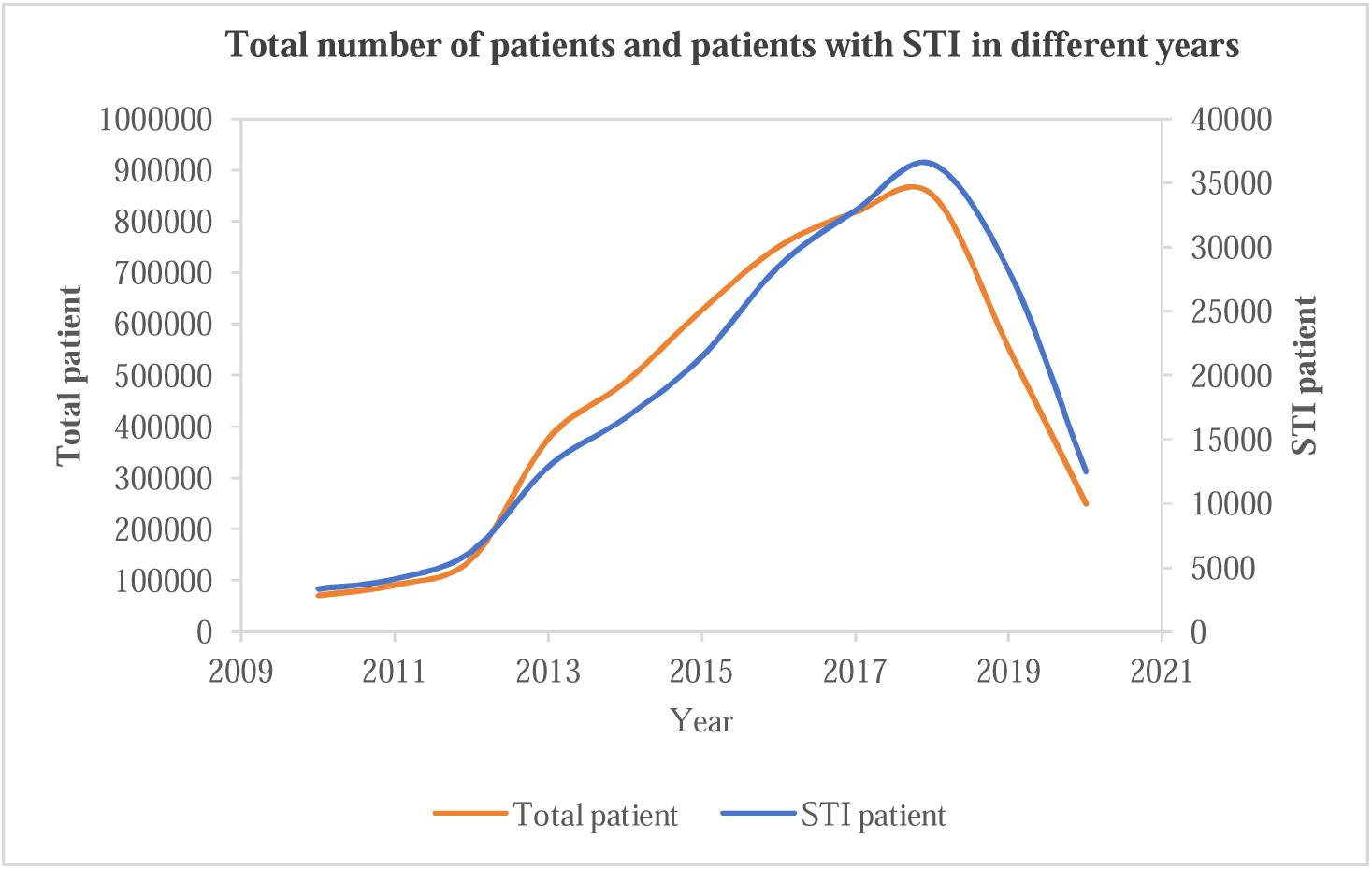
Enrolled number of patients with STI (blue) and total patients (orange) from 2010 to 2020. Both data lines increased annually starting from 2010, peaking in 2018 and then declined.

**Figure 5.**
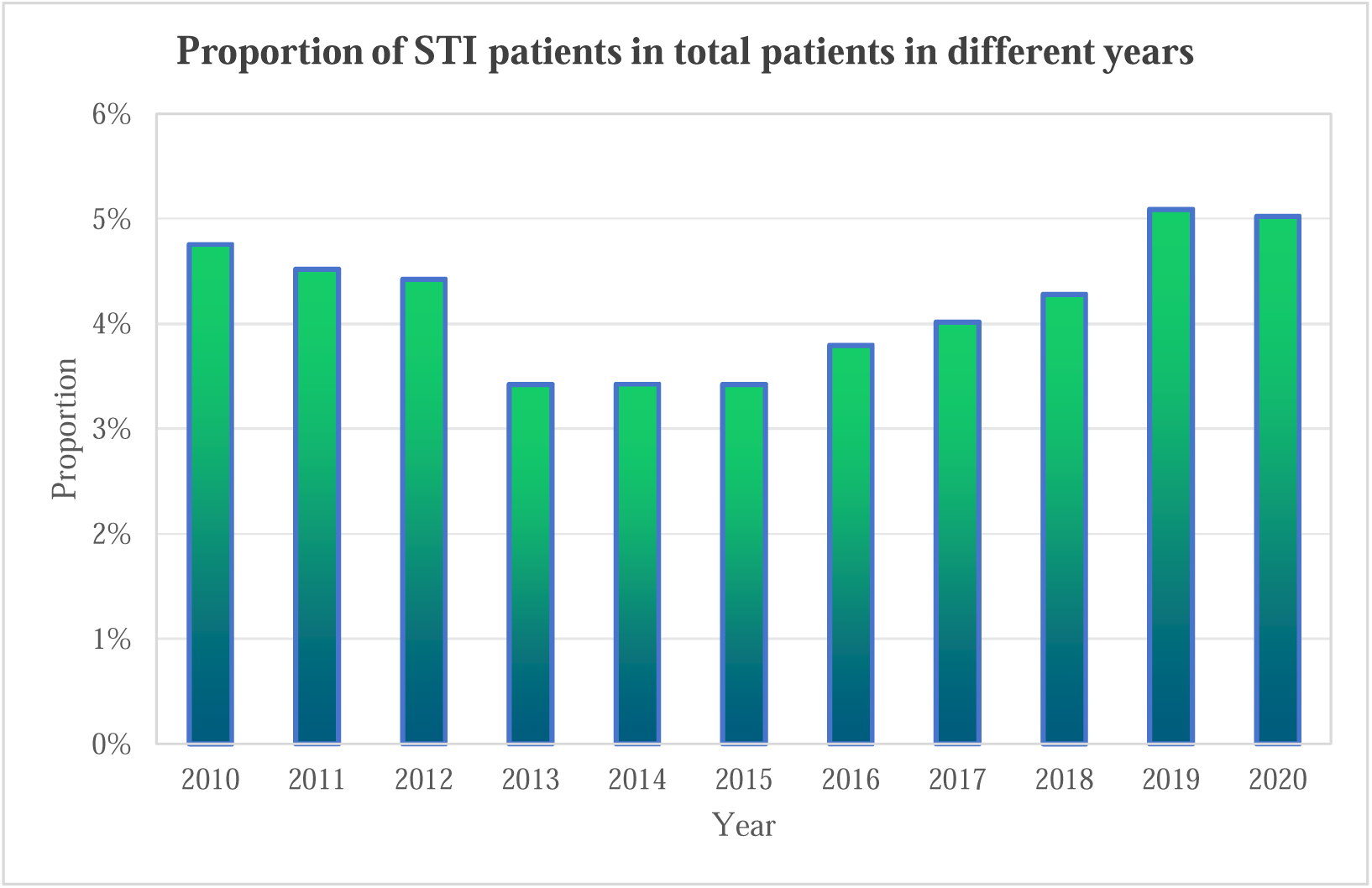
The proportion of enrolled patient with STI in total enrolled patients from 2010 to 2020. The proportion remained in a fix range, from 3% to 5%.

In figure 4, the number of total patients and the number of patients with STIs share similar patterns across a 10-year time span, indicating a relative constant proportion relationship, which is supported by figure 5. The proportion shown in table 2 and figure 5 demonstrate the reliability of our sample data.

Table 3 lists the total number of patients, the volume of identified STI information, and their corresponding proportions in each EMR sub-dataset.

**Table 3.**
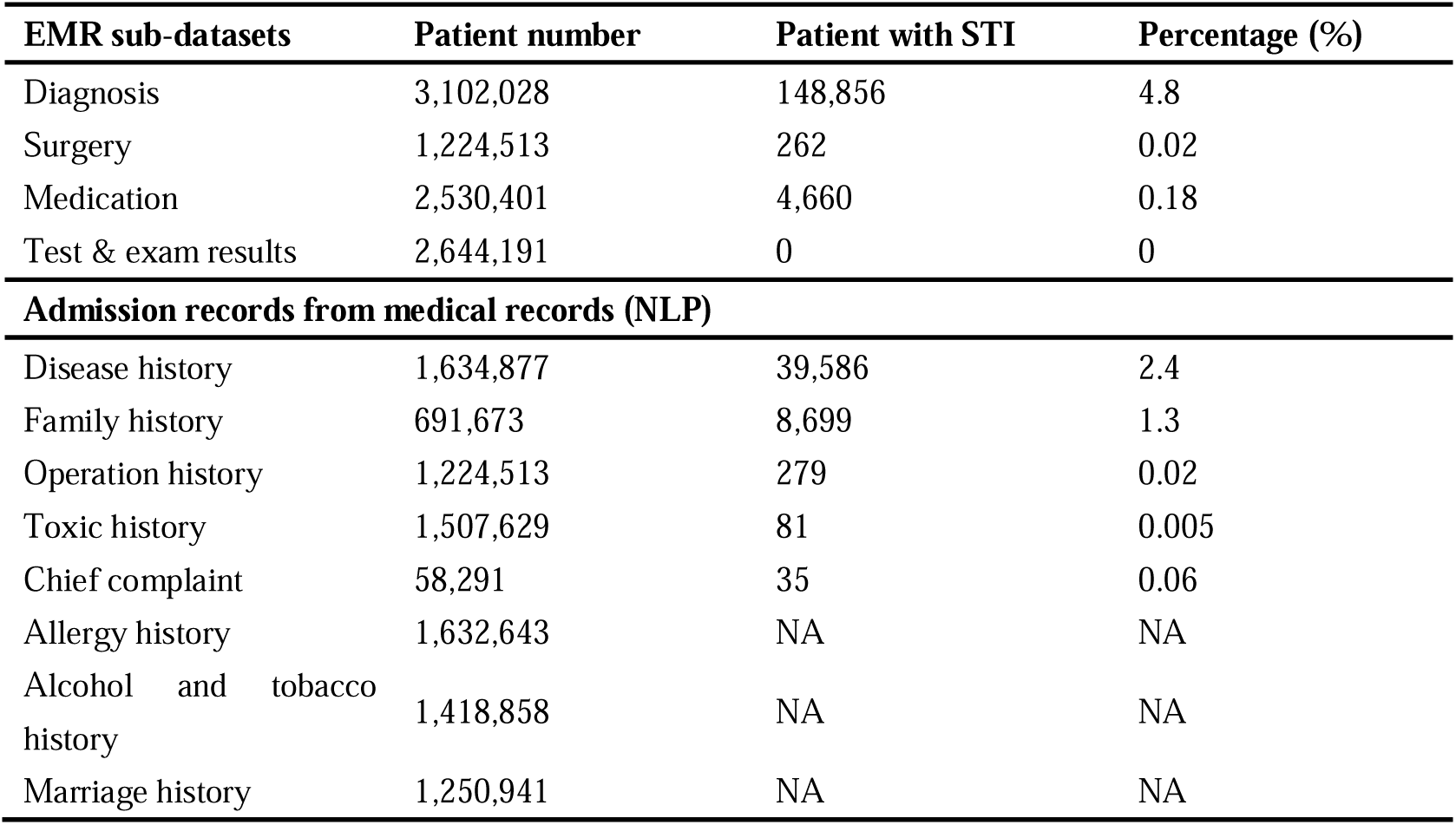
Patient identified by EPSTII. From left to right, the columns display the total number of patients, the number of identified patient number, and their corresponding proportions.

| EMR sub-datasets | Patient number | Patient with STI | Percentage (%) |
| --- | --- | --- | --- |
| Diagnosis | 3,102,028 | 148,856 | 4.8 |
| Surgery | 1,224,513 | 262 | 0.02 |
| Medication | 2,530,401 | 4,660 | 0.18 |
| Test & exam results | 2,644,191 | 0 | 0 |
| **Admission records from medical records (NLP)** |  |  |  |
| Disease history | 1,634,877 | 39,586 | 2.4 |
| Family history | 691,673 | 8,699 | 1.3 |
| Operation history | 1,224,513 | 279 | 0.02 |
| Toxic history | 1,507,629 | 81 | 0.005 |
| Chief complaint | 58,291 | 35 | 0.06 |
| Allergy history | 1,632,643 | NA | NA |
| Alcohol and tobacco history | 1,418,858 | NA | NA |
| Marriage history | 1,250,941 | NA | NA |

The number of patients with STIs identified through diagnosis was 148,856 cases, accounting for 4.8% of the patients in the diagnosis sub-dataset. The number of patients identified through disease history reached 39,586 individuals, representing 2.4% of the patients in the disease history sub-dataset. The number of patients with STI information leakage identified through chief complaints was 35 cases, making up 0.06% of the patients in the chief complaints sub-dataset.

Test and exam results contain information such as the concentration of Hepatitis B e Antigen or Treponema Pallidum Antibody, while they do not state whether patients have STI or not. Therefore, we treated them as not containing STI information.

### Frequency of Recognition

A single patient can generate multiple records in the EMR system per visit. Consequently, individual EMRs were divided into separate records based on visits, reflecting the actual EMR storage used in RWE studies. Table 4 shows the frequency of STI information identification across different sub-datasets of Chinese EMRs. As with any patient’s record, STIs can be widely identified in each sub-dataset, primarily concentrating in diagnostic records and medical record texts.

**Table 4.** Frequency of STI information identification. From left to right, the columns display the total number of records, the frequency of identified STIs, and their corresponding proportions.

| EMR sub-datasets | Visit number | STIs frequency | Percentage (%) |
| --- | --- | --- | --- |
| Diagnosis | 52,261,225 | 681,942 | 1.3 |
| Surgery | 2,275,548 | 437 | 0.02 |
| Medication | 20,602,753 | 10311 | 0.05 |
| Test & exam results | 359,784,651 | 0 | 0 |
| **Admission records from medical records (NLP)** |  |  |  |
| Disease history | 16,742,579 | 73,725 | 0.4 |
| Family history | 7,400,955 | 13,136 | 0.2 |
| Operation history | 2,275,548 | 472 | 0.02 |
| Toxic history | 13,134,334 | 91 | 0.0007 |
| Chief complaint | 188,953 | 43 | 0.02 |
| Allergy history | 4,749,858 | NA | NA |
| Alcohol and tobacco history | 3,018,618 | NA | NA |
| Marriage history | 2,089,601 | NA | NA |

In terms of privacy leakage, a total number of 681,942 STI records had been indentified from the diagnosis sub-datset with an overall recognition rate of 1.3%. In disease history records, 73,725 STIs information could be extracted from 16,742,579 records. In family history records, 13,136 STIs information could be extracted from 7,400,955 records.

### Evaluation Matrix

We selected precision and recall rates as our primary measurement criteria. During the manual curation and verification process, 1,000 records from patients with STI information and 1000 records from patients without STI information were randomly chosen from the disease history datset and reviewed by two independent medical experts to determine whether STI information had been correctly identified or ignored. After multiple trainings of our protocol, both the precision and recall rates of the EPSTII were 100%, which means that all 1000 pieces of information extracted from STI patients contained STI information, and all 1000 pieces of information extracted from non-STI patients did not contain STI information.

### Privacy Protection

After optimizing the performance of EPSTII to a satisfactory level, 2,000 complete long text entries were randomly extracted from the medical records. EPSTII was then used to identify keywords related to STIs. Under the guidance of experts, we decided to replace each identified keyword, along with 10 characters before and after it, with asterisks (*) to mask sensitive STI information and protect patient privacy. This method minimized the risk of inferring patient STI information from the EMR. Reviewers attempted to identify patients with STIs in the privacy-masked samples, and we calculated the success rate of the privacy protection strategy based on their results. In the 2,000 records with masked STI information, 10 patient was identified as carrying STI, achieving a success rate of 99.5%.

Here is one example of identifying STI information from part of medical records. {Admission diagnosis: 1. Acute Lymphoblastic Leukemia (B-ALL) 2. Hepatitis B virus carrier Diagnostic course: blood cell analysis (venous blood): white blood cell count 11.77×10^9/L, platelet count 485×10^9/L, neutrophil count 8.93×10^9/L. Hepatitis B markers: hepatitis B virus surface antigen positive (+); hepatitis B virus e antibody positive (+); hepatitis B virus core antibody positive (+). EB virus, cytomegalovirus negative. Abdominal B-Ultrasound shows slightly enlarged liver.} {Admission diagnosis: 1. Acute Lymphoblastic Leukemia (B-AL*********** carrier Diagnostic course: blood cell analysis (venous blood): white blood cell count 11.77×10^9/L, platelet count 485×10^9/L, neutrophil count 8.93×10^9/L ************** antigen positive ***********antibody positive *********** core antibody positive (+). EB virus, cytomegalovirus negative. Abdominal B-Ultrasound shows slightly enlarged liver.}

## Discussion

### Principal Findings

In analyzing the extracted STI information and the frequency of STI identification, we observed that the quantity and frequency of STI information extracted from patients’ chief complaints were significantly lower than those extracted from disease histories and family histories. This suggests that patients who are aware of their own diseases are often reluctant to disclose information about STIs to their doctors due to embarrassment and fear of a negative social image. This highlights the importance of protecting the privacy of STI information. On the other hand, patients who are unaware of their condition may not instinctively associate discomfort in their reproductive area with an STI, indicating that the negative stigma surrounding STIs is deeply ingrained in people’s minds, further emphasizing the subconscious aversion to STIs. Although the extraction proportion is even smaller in the case of toxic history, this is due to a large number of records denying the source of toxins, which have lowered the overall proportion.

After multiple optimizations, EPSTII achieved 100% accuracy and recall rates. This success is primarily attributed to our continuous refinement of the keyword list and the integration of expert opinions, online resources, and literature information in each iteration. Currently, EPSTII encompasses a total of 31 keywords related to STIs, providing comprehensive coverage of these infections.

Our privacy protection approach has achieved a 99.5% success rate, ensuring that the privacy of patients with STIs is safeguarded to the greatest extent possible. However, due to improvements of our protocol happening in admission records, which did not include medication information, we failed to consider drugs specifically targeting STIs.

Additionally, we acknowledge the potential for false positives. This issue arises because we have adopted a uniform length for privacy protection policies across different diseases, which may result in the incorrect concealment of some information and thereby limit the application scope of our model. Moreover, the sample size of 2,000 is relatively small compared to the vast amount of data in the database, contributing to our high accuracy. This limitation is due to the need for manual verification by experts for each extracted data point, which restricts our ability to extract and test a larger number of samples. In future studies, we plan to adopt a more targeted privacy protection method, taking into account drugs for treating STIs, to precisely protect private information without obscuring other crucial information about patients. Additionally, we will expand the number of validation datasets to obtain more comprehensive and realistic outcomes.

To our knowledge, this study is the first large-scale investigation into privacy leakage and STI identification in Chinese EMRs. The originality of this work can be summarized as follows:

### Exploring New Observations

Our study reveals the uneven awareness of privacy protection among EMR users. Although reliable privacy protection methods are extensively discussed in the literature, their implementation in healthcare institutions remains critical,^25,26^ especially with the inclusion of sensitive STI information.^27,28^ The 2021 Personal Information Protection Law of the People’s Republic of China clarifies the use of personal privacy information, but prior efforts to establish high standards for patient privacy protection in RWE research were minimal.^29^ This study is the first in China to use a national-level EMR database for a quantitative assessment of STI-related privacy exposure risk, aiming to enhance protection strategies and lay a foundation for future research and improved privacy protocols.

### Designing New Experiments

Chinese EMRs present unique challenges in extracting STI-related privacy information due to specific terminology and data standards. Traditional methods relying on diagnostic codes are limited by incomplete records and inconsistent documentation. Physicians may not always record STI information as a diagnosis, leading to lower recall rates and potential bias. The complexity of coding systems in Chinese EMRs further complicates patient identification. The EPSTII method addresses these limitations by providing more precise results, significantly enhancing the extraction of STI information with high accuracy from both patient-based and visit-based records.

### Contributing New Knowledge

Our results show that traditional fixed-location data masking procedures can lead to unnecessary exposure of privacy information. For example, patients’ nucleic acid test results, recorded in sub-datasets, are challenging to conceal, significantly impacting patients’ privacy. Accurate identification of STI information is crucial for comprehensive privacy protection.^30^ The EPSTII method is the first to identify STI information across the entire Chinese EMR system. It employs practical data desensitization techniques, such as data invalidation, data offset, and symmetric encryption, to prevent the misuse of private data. We have determined the optimal length of additional concealed text to retain most medical information. Quantifying the identification frequency of STI information helps researchers use EMRs wisely to avoid unnecessary privacy leakage, highlighting the richness of privacy information in EMRs. Although identification frequency alone cannot fully determine privacy leakage risk, these results underscore the importance of robust data desensitization strategies.

### Limitations

Overall, this work justifies assessing privacy leakage risks and offers a reference for effective privacy protection in Chinese EMRs. However, the study has several limitations. Firstly, the privacy risk estimated in our case study are primarily based on the EMRs of a renal disease database. Despite official guidelines for writing EMRs in China, discrepancies exist between the CRDS and other data networks in terms of data structure and operating environment. Specific protocols and variables should be optimized for broader applicability. Additionally, our privacy protection method occasionally conceals excessive information, such as the hepatitis B antibody levels of healthy patients, resulting in harmless information being hidden as well. This is because we use regular expressions to enhance speed rather than employing complex NLP techniques. In future research, we plan to explore the use of machine learning methods to better protect patient privacy.

## Conclusion

Discovering an effective and practical method for protecting privacy information in EMRs is both significant and beneficial. We have shown the feasibility of applying the EPSTII method to EMRs from 19 hospitals in various regions. We believe EPSTII can offer valuable insights for patient inclusion in any infectious disease-related research using Chinese EMRs. Our protocols, tailored specifically for the Chinese EMR system, allow for accurate and complete identification and extraction of data related to STIs, ensuring effective protection. Compared to traditional methods of including STI information, the EPSTII method provides more comprehensive results.

### What is already known on this topic

Patients suffering from sexually transmitted diseases in China are exposed to a high risk of privacy disclosure.

There is no straightforward and rapid approach to safeguard the privacy information in patient records.

### What this study adds

We have developed the first privacy protection method, EPSTII.

Our EPSTII attains 99.5% privacy protection for patients with sexually transmitted diseases.

## Data Availability

All data produced are available online at The Chinese Renal Disease Data System database

## Ethics statements

### Ethical approval

Ethical approval not required.

## Acknowledgement

This work was supported by the National Key Research and Development Program of China (2021YFC2500200) and the National Key Research and Development Program of China (2023YFC27062305). This work was supported by Nanfang Hospital, Southern Medical University. We did not use generative AI in any portion of the manuscript writing.

## Data Availability

The data that support the findings of this study are available on request from the corresponding author. Due to the sensitivity of the hospital data, it cannot be made publicly available.

